# Public health actions in response to pathogen detection in sewage: a scoping review protocol

**DOI:** 10.1101/2024.09.10.24312933

**Authors:** Maarten de Jong, Jolinda de Korne-Elenbaas, Ewout Fanoy, Gertjan Medema, Miranda de Graaf, Amrish Y. Baidjoe, Maria Prins, Maarten F. Schim van der Loeff, Joost Daams, Ana Maria de Roda Husman, Janneke C.M. Heijne

## Abstract

**Background:** Infectious disease surveillance and outbreak investigations have significantly benefited from sewage monitoring as an indicator for pathogen circulation in human populations. The use of sewage surveillance accelerated during the COVID-19 pandemic with the quantification of severe acute respiratory syndrome coronavirus 2 (SARS-CoV-2) in sewage providing predictions of SARS-CoV-2 infections and hospital admissions. A comprehensive overview how sewage monitoring can further inform local and regional public health actions proactively is needed to optimize its future use. By conducting a scoping review, we aim to provide an overview of reported public health actions as a response to sewage monitoring for pathogens.

**Methods:** This scoping review will include peer-reviewed published literature from the databases MEDLINE, EMBASE and Web of Science. Literature describing public health actions as a response to sewage monitoring in the field of human infectious diseases will be considered for inclusion. Literature not written in English, published prior to 1 January 2014, systematic reviews, editorials and letters to the editor will be excluded. Screening of literature against the inclusion criteria and the subsequent data extraction will be performed by two reviewers. The described public health actions, and corresponding sewage sampling methods and microbiological analytic tools and techniques that can be applied on sewage samples for detecting pathogens will also be extracted. The extracted data from included literature will be combined into a narrative synthesis.

## Background

In the field of infectious disease surveillance, monitoring and control, innovative approaches are continuously sought to enhance our ability to detect and respond to potential outbreaks that jeopardize public health (1-4). Conventional surveillance and monitoring methods predominantly rely on clinical data, such as syndromes and laboratory test results. These are often characterized by delays in reporting, underdiagnosis, and limitations in coverage.

Sewage, as a collective indication of a community’s health, provides a dynamic and real-time reservoir of biological information, offering a more timely and potentially broad view on the population’s health status. Therefore, sewage monitoring can complement other surveillance tools by tracking, analyzing and responding to the presence of various pathogen substances such as genomic material of pathogens in sewage (5-7).

Sewage monitoring has been used for decades to track the spread of polio, norovirus, hepatitis E, and various other pathogens in communities (8-10). The use and acceptance of sewage monitoring has led to contributions to public health actions such as targeted monitoring of infectious diseases on neighborhood, institutional and community level, and the guidance of vaccination efforts (11-13). In recent years, the COVID-19 pandemic accelerated developments in the field of sewage monitoring globally as it proved to be a cost-effective, rapid and reliable source of information on the spread of severe acute respiratory syndrome coronavirus 2 (SARS-CoV-2) and its variants in the population (14-17). As a result, numerous high-income countries developed sewage monitoring infrastructure for proactive surveillance and response to future infectious disease outbreaks. The opportunity to apply sewage monitoring in low- and middle-income countries is also being explored and first steps have been taken (18-20).

Scoping reviews are a useful tool to map out the existing literature on a specific topic by identifying key concepts and research gaps. Various recent reviews in the fields of sewage monitoring of pathogens have provided comprehensive insights into the practical and technical use of sewage monitoring for outbreak management and surveillance (16, 17, 21). One review described the potential of sewage surveillance to guide public health response or policy prior to the SARS-CoV-2 pandemic (22). However, an extensive and detailed overview of public health actions as a response to pathogen detection in sewage with corresponding sewage monitoring strategies has to our knowledge not yet been created.

### Objective of the review

This scoping review aims to assess the potential of sewage monitoring in infectious disease control by providing an overview of public health actions taken in response to pathogen detection in sewage and their corresponding sewage sampling strategies and sewage analysis methods (Figure 1). The review intends to offer guidance to public health professionals on how public health actions can be effectively informed by sewage monitoring.

**Figure 1.**
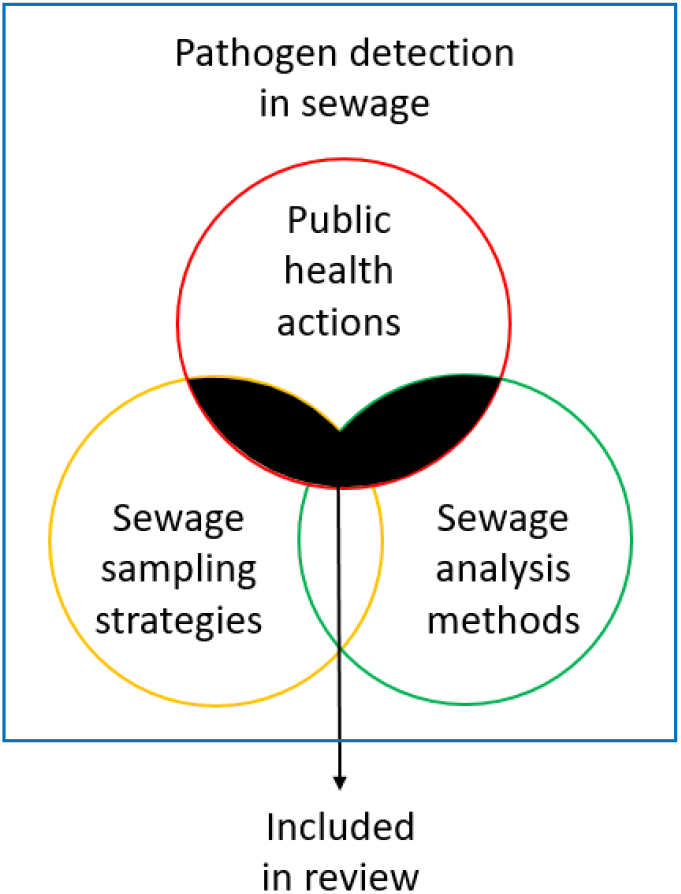
Literature on public health actions as a response to sewage sampling strategies and sewage analysis methods used for pathogen detection in sewage included in this review

Specific sub-objectives:

1. To identify public health actions as a response to pathogen detection in sewage;
2. To identify sewage sampling strategies for pathogen detection resulting in public health actions;
3. To identify analysis methods for pathogens in sewage samples resulting in public health actions.

We define ‘pathogen detection in sewage’ as the process of identifying genomic material of pathogens (bacteria, viruses, fungi and parasites) in sewage. We define ‘public health actions’ as the efforts aimed at infectious disease management and control, including response and surveillance, tailored interventions, targeted testing, contact tracing, transmission analyses, etc. As a guideline, we used the Essential Public Health Functions composed by the World Health Organization (WHO) to contextualize public health actions for the focus of this scoping review (23). We define ‘sewage’ as untreated wastewater containing a mixture of human waste, domestic and industrial wastewater, and other debris. ‘Sewage sampling’ is defined as the process of collecting sewage samples from one or more locations for analysis and testing purposes. We define ‘sewage analysis’ as the examination and assessment of the composition and characteristics of sewage to identify genomic material of pathogens, such as bacteria, viruses, fungi and parasites.

## Methods

### Searching for articles

The search strategy aims to identify peer-reviewed published literature with a focus on public health, sewage sampling methods and related analytic tools and techniques in the field of infectious diseases that can be applied on sewage samples. The databases MEDLINE, Embase, and Web of Science will be searched for literature using searches developed in collaboration with an information specialist (JD) from the Amsterdam UMC library. Searches will be conducted on title, abstract and keywords and search language will be restricted to English. Once consensus is reached regarding relevance of the articles full searches will be run in all databases. See Appendix 1 for the detailed search string.

### Screening process

All peer-reviewed literature results will be imported to and managed with Rayyan software, a web-based systematic review tool that assists in expediting the screening phase (24). Duplicates will be removed. Before commencing screening, consistency checking on in- and exclusion will be performed on a subset of records at both title and abstract levels. Initially, titles and abstracts of a random sample of 300 articles are assessed independently by two reviewers (MJ and JK) according to the inclusion criteria. The results will then be compared and all discrepancies will be discussed. Subsequently, the remaining literature will be screened by the two reviewers and the results will be compared and discussed after 25%, 50%, 75% and 100% of the literature has been screened. In the next stage, a full-text review is conducted of the included literature to determine eligibility. In both stages, the reviewers work independently and subsequently will compare the results. Any discrepancies will be resolved through consensus, and if that is not reached, by a third senior reviewer (JH). The review team will ensure that reviewers who have authored articles to be considered within the review have no role in decisions regarding the inclusion or data coding of their own work. Literature not meeting the inclusion criteria will be excluded. The final search results will be reported in a PRISMA flow diagram (25).

### Eligibility criteria

Literature with no explicit or direct link to public health actions will be excluded, as will be literature dealing with sewage sampling and sewage analysis outside the field of human infectious diseases. All literature not written in English, non-peer reviewed literature, and all literature published before 1 January 2014 will be excluded since the field of sewage monitoring has developed significantly in the recent decade. Also systematic reviews, commentaries, editorials and letters to the editor will be excluded.

### Study validity assessment

Formal quality assessment of the included literature will not be undertaken, as the aim of this scoping review is to summarize and interpret public health actions reported in the literature, rather than to assess or compare the methodological quality or calculated measurements of the included literature (26).

### Data coding and extraction strategy

A form for data extraction and characterization will be used and will include: study title, first author(s), other author(s), publication year, journal, calendar year(s) of data collection, geographical context, aim(s) description, targeted pathogen(s), target population, clinical data, commissioner, end user, sampling naming, sampling method(s), sampling location(s), sampling technique, analysis naming, type of analysis, analysis technique(s), analysis target, sample processing methodology, data normalization, analysis protocol(s), outcome measure(s), description public health action(s), public health action(s) as a response to the conducted sewage monitoring and recommendations (Table 1). The form will be pre-tested and adapted as needed. Two reviewers will extract data independently. Extracted data will be compared and reviewers (MJ and JK) will discuss discrepancies in the data extraction until consensus is reached. Unresolved discrepancies will be arbitrated by two additional reviewers (JH and AMRH). Authors of articles will be consulted for additional information and clarification where necessary during the extraction process. The data extraction form will be refined during the review process and based on the emerging themes from the literature. Extracted data records will be made available as additional files in the final report.

**Table 1.**
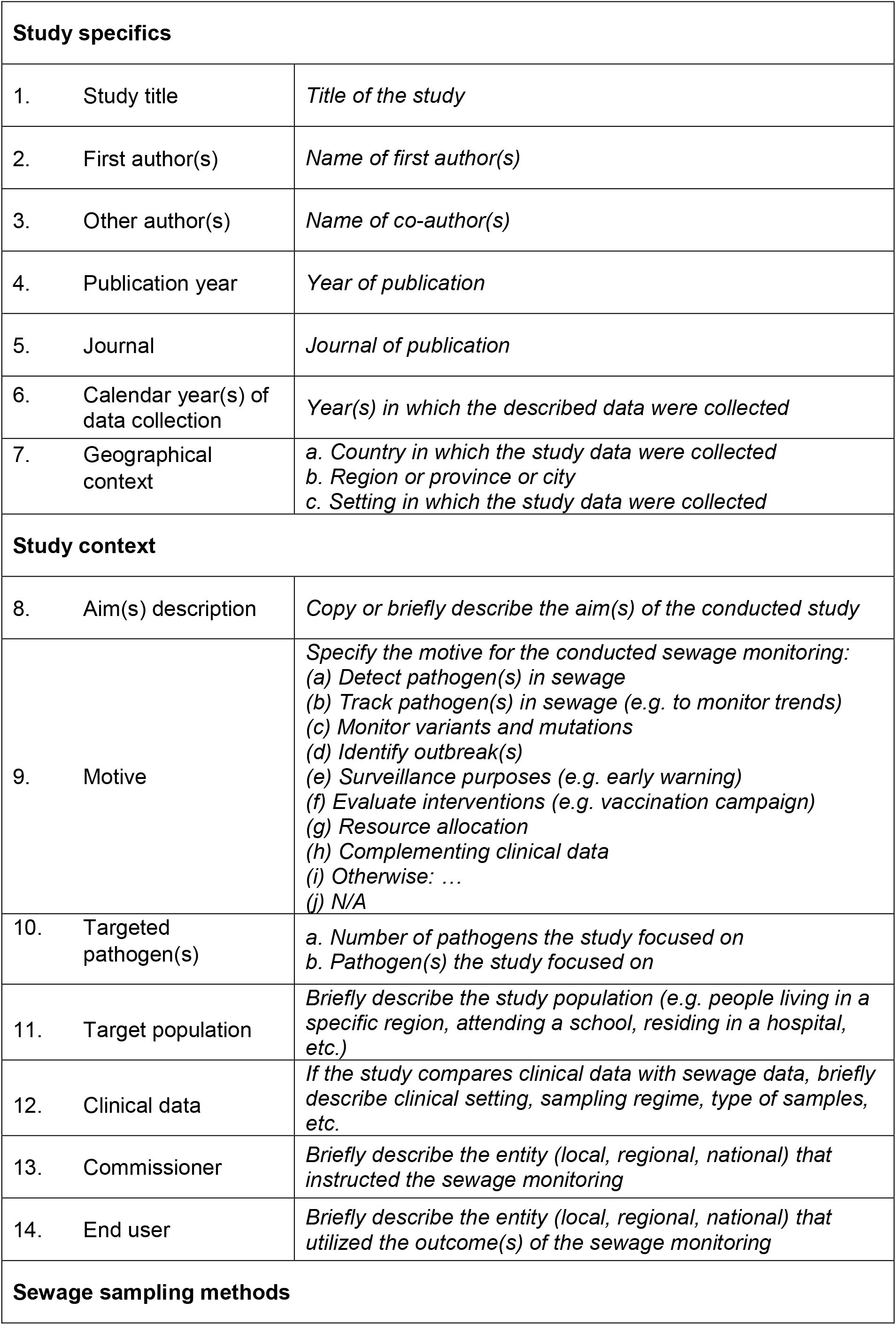
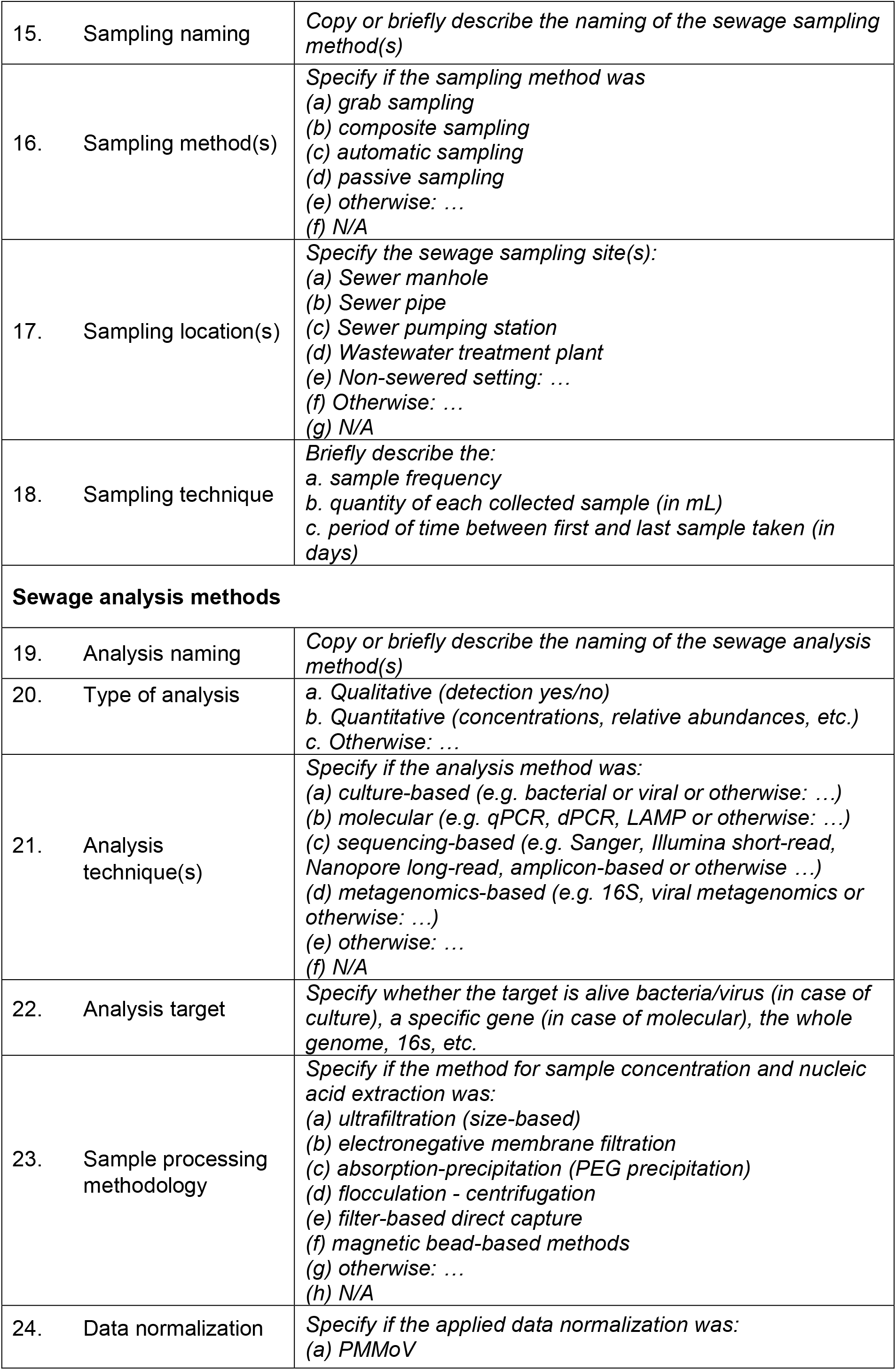
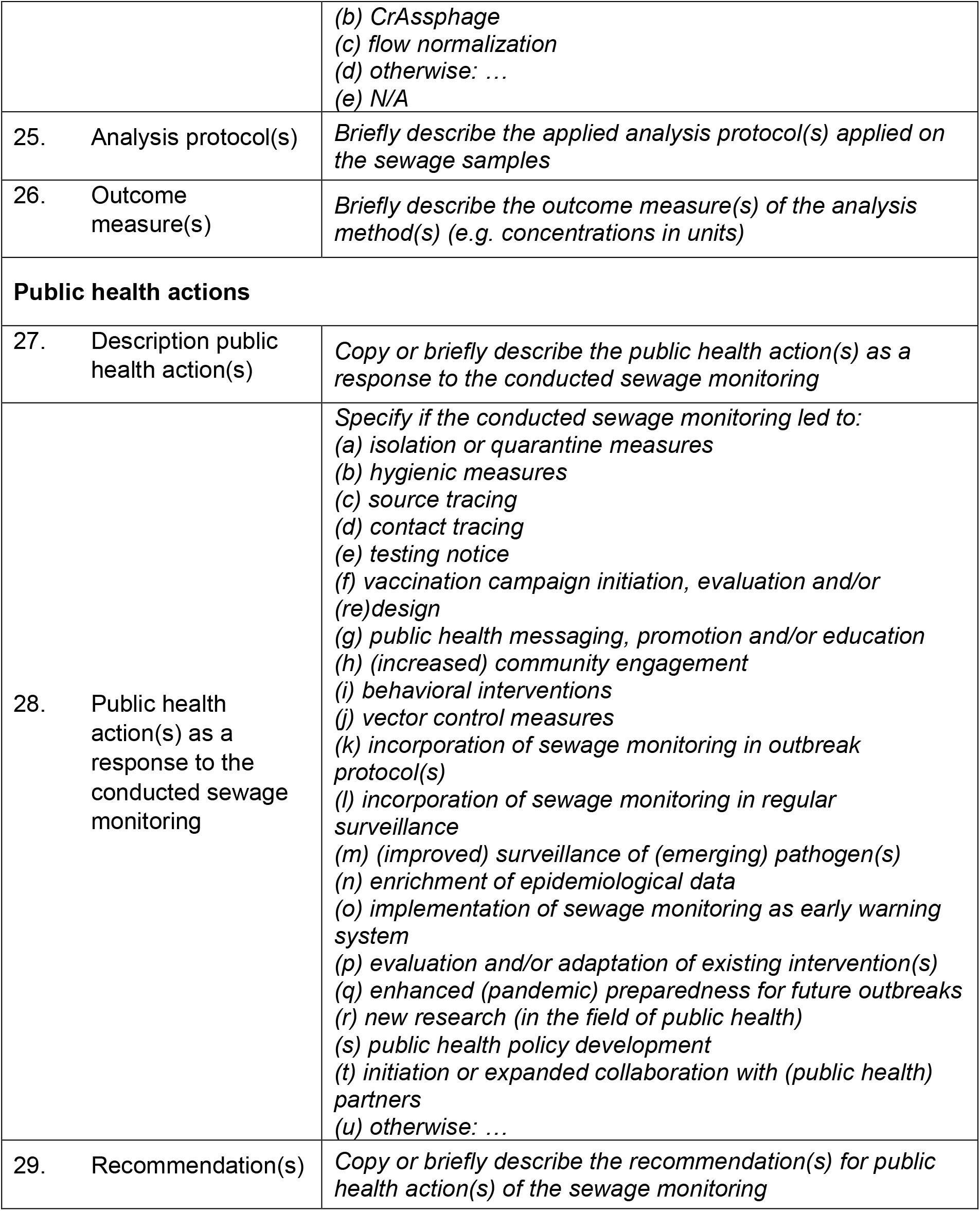
Data extraction sheet.

|  |  |
| --- | --- |
| **Study specifics** |  |
| 1. Study title | *Title of the study* |
| 2. First author(s) | *Name of first author(s)* |
| 3. Other author(s) | *Name of co-author(s)* |
| 4. Publication year | *Year of publication* |
| 5. Journal | *Journal of publication* |
| 6. Calendar year(s) of data collection | *Year(s) in which the described data were collected* |
| 7. Geographical context | *a. Country in which the study data were collected*<br>*b. Region or province or city*<br>*c. Setting in which the study data were collected* |
| **Study context** |  |
| 8. Aim(s) description | *Copy or briefly describe the aim(s) of the conducted study* |
| 9. Motive | *Specify the motive for the conducted sewage monitoring:*<br>*(a) Detect pathogen(s) in sewage*<br>*(b) Track pathogen(s) in sewage (e.g. to monitor trends)*<br>*(c) Monitor variants and mutations*<br>*(d) Identify outbreak(s)*<br>*(e) Surveillance purposes (e.g. early warning)*<br>*(f) Evaluate interventions (e.g. vaccination campaign)*<br>*(g) Resource allocation*<br>*(h) Complementing clinical data*<br>*(i) Otherwise: ...*<br>*(j) N/A* |
| 10. Targeted pathogen(s) | *a. Number of pathogens the study focused on*<br>*b. Pathogen(s) the study focused on* |
| 11. Target population | *Briefly describe the study population (e.g. people living in a specific region, attending a school, residing in a hospital, etc.)* |
| 12. Clinical data | *If the study compares clinical data with sewage data, briefly describe clinical setting, sampling regime, type of samples, etc.* |
| 13. Commissioner | *Briefly describe the entity (local, regional, national) that instructed the sewage monitoring* |
| 14. End user | *Briefly describe the entity (local, regional, national) that utilized the outcome(s) of the sewage monitoring* |
| **Sewage sampling methods** |  |

|  |  |  |
| --- | --- | --- |
| 15. | Sampling naming | *Copy or briefly describe the naming of the sewage sampling method(s)* |
| 16. | Sampling method(s) | *Specify if the sampling method was*<br>*(a) grab sampling*<br>*(b) composite sampling*<br>*(c) automatic sampling*<br>*(d) passive sampling*<br>*(e) otherwise: ...*<br>*(f) N/A* |
| 17. | Sampling location(s) | *Specify the sewage sampling site(s):*<br>*(a) Sewer manhole*<br>*(b) Sewer pipe*<br>*(c) Sewer pumping station*<br>*(d) Wastewater treatment plant*<br>*(e) Non-sewered setting: ...*<br>*(f) Otherwise: ...*<br>*(g) N/A* |
| 18. | Sampling technique | *Briefly describe the:*<br>*a. sample frequency*<br>*b. quantity of each collected sample (in mL)*<br>*c. period of time between first and last sample taken (in days)* |
| **Sewage analysis methods** |  |  |
| 19. | Analysis naming | *Copy or briefly describe the naming of the sewage analysis method(s)* |
| 20. | Type of analysis | *a. Qualitative (detection yes/no)*<br>*b. Quantitative (concentrations, relative abundances, etc.)*<br>*c. Otherwise: ...* |
| 21. | Analysis technique(s) | *Specify if the analysis method was:*<br>*(a) culture-based (e.g. bacterial or viral or otherwise: ...)*<br>*(b) molecular (e.g. qPCR, dPCR, LAMP or otherwise: ...)*<br>*(c) sequencing-based (e.g. Sanger, Illumina short-read, Nanopore long-read, amplicon-based or otherwise: ...)*<br>*(d) metagenomics-based (e.g. 16S, viral metagenomics or otherwise: ...)*<br>*(e) otherwise: ...*<br>*(f) N/A* |
| 22. | Analysis target | *Specify whether the target is alive bacteria/virus (in case of culture), a specific gene (in case of molecular), the whole genome, 16s, etc.* |
| 23. | Sample processing methodology | *Specify if the method for sample concentration and nucleic acid extraction was:*<br>*(a) ultrafiltration (size-based)*<br>*(b) electronegative membrane filtration*<br>*(c) absorption-precipitation (PEG precipitation)*<br>*(d) flocculation - centrifugation*<br>*(e) filter-based direct capture*<br>*(f) magnetic bead-based methods*<br>*(g) otherwise: ...*<br>*(h) N/A* |
| 24. | Data normalization | *Specify if the applied data normalization was:*<br>*(a) PMMoV* |
|  |  | **(b) CrAssphage**<br>**(c) flow normalization**<br>**(d) otherwise: ...**<br>**(e) N/A** |
| 25. | Analysis protocol(s) | *Briefly describe the applied analysis protocol(s) applied on the sewage samples* |
| 26. | Outcome measure(s) | *Briefly describe the outcome measure(s) of the analysis method(s) (e.g. concentrations in units)* |
| **Public health actions** |  |  |
| 27. | Description public health action(s) | *Copy or briefly describe the public health action(s) as a response to the conducted sewage monitoring* |
| 28. | Public health action(s) as a response to the conducted sewage monitoring | *Specify if the conducted sewage monitoring led to:*<br>**(a) isolation or quarantine measures**<br>**(b) hygienic measures**<br>**(c) source tracing**<br>**(d) contact tracing**<br>**(e) testing notice**<br>**(f) vaccination campaign initiation, evaluation and/or (re)design**<br>**(g) public health messaging, promotion and/or education**<br>**(h) (increased) community engagement**<br>**(i) behavioral interventions**<br>**(j) vector control measures**<br>**(k) incorporation of sewage monitoring in outbreak protocol(s)**<br>**(l) incorporation of sewage monitoring in regular surveillance**<br>**(m) (improved) surveillance of (emerging) pathogen(s)**<br>**(n) enrichment of epidemiological data**<br>**(o) implementation of sewage monitoring as early warning system**<br>**(p) evaluation and/or adaptation of existing intervention(s)**<br>**(q) enhanced (pandemic) preparedness for future outbreaks**<br>**(r) new research (in the field of public health)**<br>**(s) public health policy development**<br>**(t) initiation or expanded collaboration with (public health) partners**<br>**(u) otherwise: ...** |
| 29. | Recommendation(s) | *Copy or briefly describe the recommendation(s) for public health action(s) of the sewage monitoring* |

### Potential effect modifiers/reasons for heterogeneity

Not applicable.

### Data synthesis and presentation

The data will be aggregated in an Excel sheet for validation and coding. Each row will represent a study, each column a data item to be extracted and the cells will contain information gathered from the selected literature. The review will employ narrative synthesis methods and show descriptive statistics, tables, and figures that summarize the evidence base. The descriptive table will include data extracted (as mentioned above) of included studies. The textual description will be thematically categorized to answer specific objective 1 by identifying public health actions as a response to pathogen detection in sewage. To answer specific objective 2, we will categorically summarize sewage sampling strategies for pathogen detection for specific public health actions. To answer specific objective 3, we will categorically summarize analysis methods for pathogens in sewage samples for specific public health actions. Finally, we will provide a summary of the diverse public health actions as a response to pathogen detection in sewage and describe suitable sewage sampling and analysis methods.

## Data Availability

All data produced in the present study are available upon reasonable request to the authors

## Declarations

### Ethics approval and consent to participate

Not applicable.

### Consent for publication

Not applicable.

### Availability of data and materials

The datasets generated and/or analyzed during the current study will become available in the Dataverse repository (27).

### Authors’ contribution

MJ, AMRH, JH, MS and JK designed the study. JD advised about the design of the scoping review. JD and MJ developed the search strategy. EF and MP delivered expert opinion on the study design. MJ drafted the manuscript of this protocol. All authors contributed to the protocol and all approved the final version.

## Acknowledgements

The authors acknowledge the research infrastructure provided by the Dutch Collaborative Academic practice for Public health Infectious diseases (CAPI).

## Appendix 1

### Ovid Em base Classic+Embase

#### Searches

Sewage/

(exp wastewater/ or sewage/ or exp water pollution/) and (exp communicable disease control/ or exp epidemiological monitoring/ or environmental

monitoring/ or exp environmental surveillance/ or water monitoring/)

Wastewater-Based Epidemiology/

((wastewater? or sewage) adj3 (sampl*or surveil*or monitor* or treatment plant? or test*)).ab,kw,ti.

((wastewater? or sewage) adj10 (detect* or charact* or concentration)).ab,kw,ti.

wastewater microbiome.ab,kw,ti.

sewage signal*.ab,kw,ti.

or/1-7 [sewage sampling]

exp communicable disease control/ or public health/

epidemiology.fs.

(public health or lockdown or virus spread or epidemiol* or epidemic* or endemic* or pandemic* or community wastewater? or ((spread or surveillance)

adj5 (infection or disease or virus* or norovirus*or bacter* or parasit* or fung*)) or (spread adj3 infection)).ab,kw,ti.

or/9-11 [public health]

exp *infection/ or *fungus/ or *parasite/ or *virus/

(communicable disease? or fungi or bacter* or parasite? or virus* or viral or norovirus or infect*).ab,kw,ti.

8 and 12

limit 15 to covid-19

or/13-14,16 [infectious agents]

and/8,12,17

wastewater based epidemiology.mp.

18 or 19

**Ovid M EDLINE(R)**

#### Searches

Sewage/ch, is, ip, mt, mi, ps, pc, vi

(wastewater/ or sewage/ or water pollution/) and (exp communicable disease control/ or epidemiological monitoring/ or exp environmental monitoring/)

(wastewater/ or sewage/ or water pollut ion/) and isolation purification.fs.

Wastewater-Based Epidemiological Monitoring/

((wastewater? or sewage) adj3 (sampl* or surveil* or monitor* or treatment plant? or test*)).ab,kf,ti.

((wastewater? or sewage) adj10 (detect * or charact* or concentration)).ab,kf,ti.

wastewater microbiome.ab,kf,ti.

sewage signal*.ab,kf,ti.

or/1-8 I sewage sampling)

communicable disease control/ or public health/

epidemiology.ls.

(public health or lockdown or virus spread or epidemiol* or epidemic* or endemic* or pandemic* or community wastewater? or ((spread or surveillance)

adj5 (infection or disease or virus* or norovirus* or bacter* or parasit* or fung*)) or (spread adj3 infection)).ab,kf,ti.

or/10-12 [public health]

communicable diseases/

(communicable disease? or fungi or bacter* or parasite? or virus* or viral or norovirus or infect*).ab,kf,ti.

9 and 13

limit 16 to covid-19

or/14-15,17 [infectious agents]

and/9,13,18

wastewater based epidemiological monitoring/

or 20

**Web of Science Core Collection**

#### Searches

**#**

TS=((wastewater? or sewage) NEAR/2 (sampl* or surveil’ or monitor* or “treatment plant?” or test*))

TS=((wastewater? or sewage) NEAR/9 (detect* or charact* or concentration))

TS=“wastewater microbiome”

TS=“sewage signal*”

#4 OR #3 OR #2 OR #1

TS=(“public health” or lockdown or virus spread or epidemiol* or epidemic* or endemic* or pandemic* or “community wastewater?” or ((spread or

surveillance) NEAR/4 (infection or disease or virus* or norovirus* or bacter* or parasit* or fung*)) or (spread NEAR/2 infect*)

#5 AND #6 AND #7

